# Circulating cytokines and lymphocyte subsets in patients who have recovered from COVID-19

**DOI:** 10.1101/2020.07.22.20160259

**Authors:** Hasichaolu, Xinri Zhang, Xin Li, Xin Li, Dongyan Li

## Abstract

To investigate the immune status of people who previously had COVID-19 infections, we recruited patients 2 weeks post-recovery and analyzed circulating cytokines and lymphocyte subsets. We measured levels of total lymphocytes, CD4^+^ T cells, CD8^+^ T cells, CD19^+^ B cells, CD56^+^ NK cells, and the serum concentrations of interleukin (IL)-1, IL-4, IL-6, IL-8, IL-10, transforming growth factor beta (TGF-β), tumor necrosis factor alpha (TNF-α), and interferon gamma (IFN-γ) by flow cytometry. We found that in most post-recovery patients, levels of total lymphocytes (66.67%), CD3^+^ T cells (54.55%), CD4^+^ T cells (54.55%), CD8 ^+^ T cells (81.82%), CD19^+^ B cells (69.70%), and CD56^+^ NK cells(51.52%) remained lower than normal, whereas most patients showed normal levels of IL-2 (100%), IL-4 (80.88%), IL-6 (79.41%), IL-10 (98.53%), TNF-α (89.71%), IFN-γ (100%) and IL-17 (97.06%). Compared to healthy controls, 2-week post-recovery patients had significantly lower absolute numbers of total lymphocytes, CD3^+^ T cells, CD4^+^ T cells, CD8^+^ T cells, CD19^+^ B cells, and CD56^+^ NK cells, along with significantly higher levels of IL-2, IL-4, IL-6, IL-10, TNF-α, IFN-γ and IL-17. Among post-recovery patients, T cells, particularly CD4^+^ T cells, were positively correlated with CD19^+^ B cell counts. Additionally, CD8^+^ T cells positively correlated with CD4^+^ T cells and IL-2 levels, and IL-6 positively correlated with TNF-α and IFN-γ. These correlations were not observed in healthy controls. By ROC curve analysis, post-recovery decreases in lymphocyte subsets and increases in cytokines were identified as independent predictors of rehabilitation efficacy. These findings indicate that the immune system has gradually recovered following COVID-19 infection; however, the sustained hyper-inflammatory response for more than 14 days suggests a need to continue medical observation following discharge from the hospital. Longitudinal studies of a larger cohort of recovered patients are needed to fully understand the consequences of the infection.

## 1. Introduction

Since December 2019, severe cases of pneumonia have been reported in Wuhan, Hubei Province, China, and the disease has since spread rapidly throughout China and to overseas countries [1]. Several days later, the virus was identified as a new *Betacoronavirus*, which was officially named severe acute respiratory syndrome coronavirus 2 (SARS-CoV-2), with the accompanying pneumonia referred to as coronavirus disease 2019 (COVID-19) [2]. By taking a series of preventive control and medical treatment measures, the rise of the epidemic within China has been contained to a certain extent [3]. In mid-April 2020, the WHO had announced that approximately 1,914,916 people were infected with the disease globally, with 123,010 deaths, and 501,758 people having recovered [4]. Although patients can recover from the infection, some of the side effects may have a significant impact on recovered patients in the future. It is therefore imperative to understand possible outcomes of COVID-19 recovered patients and determine if they are at higher risk of other detrimental illnesses using longitudinal analyses. Moreover, it is necessary to follow-up with these recovered patients and perform comprehensive assessments for appropriate management of their physical and psychological health [4].

Upon SARS-CoV-2 infection, the immune system carries out an immune response to defend against the virus. At the present, a series of studies regarding the immune status and prognosis of patients with COVID-19 have been done [5,6]. Some of these studies focused on profiling cytokines in the peripheral blood of hospitalized patients with COVID-19, with one study reporting a significant increase in IL-6 and IL-10 in patients with severe cases of COVID-19 [5]. However, the long-term consequences in patients who recovered and were discharged are still unknown. Lymphocytes play a critical role in maintaining immune function. Recent studies have observed that levels of lymphocytes in patients with COVID-19 often decrease [5], but characteristics of lymphocyte subsets in patients who recover remain unknown. In this study, we aimed to detect lymphocyte subsets and cytokines in peripheral blood from patients who have recovered from COVID-19 to explore the immune status of these patients. This may have clinical value, by aiding in retrospective diagnoses and prognostic assessments of patients during the rehabilitation period, and to provide a quantitative basis for intervention.

## 2. Materials and Methods

### 2.1. Materials

From February 2020 to March 2020, we enrolled a total of 69 patients who were previously infected with SARS-CoV-2. These patients had been confirmed to be positive for SARS-CoV-2 nucleic acid by real-time fluorescent RT-PCR. All patients had been treated in hospital, then discharged when physical indices were in line with the discharge standard based on the national “Diagnosis and Treatment Protocol for Novel Coronavirus Pneumonia (Trial Version 7)” [7]. After 2 weeks of isolation, these patients were regarded as completely recovered, with no SARS-CoV-2 remaining in the body and a negative nucleic acid detection test.

In addition, we recruited subjects who had undergone physical examination in the Health Examination Center of The First Hospital of Shanxi Medical College during April 2020. These subjects were used as healthy controls (HCs) from whom serum samples were collected after obtaining informed consent. All recruited subjects including 2-week post-recovery patients and healthy controls completed a questionnaire including questions regarding diseases, prescribed and over-the-counter medication, presence of fever, allergy and eczema, and a general question concerning subjective health. Demographic data included diet, exercise status, ethnicity, and body mass index. Patients were excluded if they had a history of illness including acute or chronic infection, hepatobiliary diseases, hematological diseases, urinary system diseases, nutrition and metabolism diseases, rheumatic diseases, endocrine diseases, circulatory system diseases, muscle trauma, hypertension, obesity (weight is 10% more than the average of the reference population with the same sex and height), or malnutrition. Further, the included patients would be excluded if they fulfilled any of the following testing criteria: positive in HBsAg, HCV antibody, or HIV antibody; creatinine above 120 μmol/L, creatine kinase above 500 U/L, uric acid above 475 μmol/L, glucose above 7.0 mmol/L, or C-reactive protein above 12.0 mg/L. In total, 68 recovered patients, including 48 males, aged 21-49 years-old, and 20 females, aged 24-66 years-old, along with 28 healthy controls, including 8 males, aged 21-53 years-old, and 20 females, aged 22-53 years-old were included.

This study was approved by the ethics committee of The First Hospital of Shanxi Medical University. Written informed consent was obtained from all patients.

### 2.2. Reagents

Cytokine detection reagents were provided by Jiangxi Saiji Biotechnology Limited Company (calibrator lot number: 20190801). Lymphocyte subset detection kits were from Beckman Coulter Incorporated (USA).

### 2.3. Sample Collection

2 mL of venous blood was collected from all subjects into EDTA anticoagulation tubes. 4 mL of venous blood was collected into gold-topped serum-separating tubes with separating gel, then centrifuged at 2000-3000 rpm for 5 min to obtain serum. Samples exhibiting hemolysis, jaundice or high lipid levels were removed.

### 2.4. Lymphocyte Subset Detection

Two flow tubes were prepared for each sample and labeled A and B, respectively. 10 μL of each monoclonal antibody were added to tube A (CD45-FITC/CD4-RD1/ CD8-ECD/CD3-PC5 antibody) or tube B (CD45-FITC/CD56-RD1/ CD19-ECD/CD3-PC5 antibody). 100 μL of EDTA-anticoagulated whole blood were then added to the corresponding flow tubes, and the tubes were vortexed and incubated at room temperature for 20 min in the dark. Next, 1 mL of lysing solution was added to all tubes. After incubating tubes at room temperature for an additional 20 min in the dark, samples were centrifuged at 200 x *g* in a low-speed centrifuge for 5 min, then supernatant was aspirated, and 2 mL of PBS were added to each tube. Samples were centrifuged again at 200 x *g* for 5 min, supernatant was aspirated, and 500 μL of PBS were added. The samples were then analyzed by fluorescence-labeled flow cytometry on a DxFLEX flow cytometer (Beckman Coulter).

### 2.5. Cytokine Detection

Venous blood samples were collected into tubes containing separating gel and centrifuged at 2000-4000rpm for 20 min to obtain serum for analysis. Seven cytokines including IL-2, IL-4, IL-6, IL-10, IL-17, TNF-α, and IFN-γ were detected by multiple microsphere flow immunofluorescence according to the manufacturer’s instructions. After blood samples and the corresponding flow tube were numbered from 1 to 27, the captured microsphere mixture was centrifuged at 200 x *g* in a low-speed centrifuge for 5 min, and supernatant was carefully aspirated. Microsphere buffer of the same volume as the supernatant was then added, and samples were mixed on a whirlpool well then incubated in the dark for 30 min. Next, 25 μL of the above incubated captured microsphere mixture, 25 μL of centrifuged serum, and 25 μL of fluorescent reagents were added to the corresponding flow tube, and all tubes were mixed well. After incubating at room temperature for 2.5 hours in the dark, 1 mL of PBS was added into each flow tube. After centrifuging at 200 x *g* for 5 min, the supernatant was carefully aspirated, and 100 μL of PBS were added to each flow tube. Fluorescent detection was then performed on a calibrated flow cytometer for each sample in sequence.

### 2.6. Statistics

All statistical analyses were performed using EXCEL (Microsoft) and SPSS Statistics version 21.0 software. Briefly, the data were inspected using scatter and distribution plots, outliers were removed following Dixon’s rule, and normality was determined using the Shapiro-Wilk test. Continuous variables were expressed as x ± s, and the comparison between groups was analyzed by t-tests. Non-continuous variables were expressed as median with interquartile range (IQR), and the comparison between groups was analyzed using nonparametric comparative tests. Correlation analysis results were expressed by Pearson or Spearman correlation coefficient, with a larger r indicating a stronger linear correlation. *P*<0.05 was considered as statistically significant. Receiver operating characteristic (ROC) curve analyses were conducted to evaluate the probability of cytokines and lymphocyte subsets in predicting recovery efficacy.

## 3. Results

### 3.1. Baseline Data

The study population included 68 patients 2 weeks after recovering from COVID-19 (48 males and 20 females) with a mean age of 33.98 ± 8.03 (21-66) years-old, and 28 healthy controls (8 males and 20 females) with a mean age of 35.13 ± 8.85 (21-49) years-old.

### 3.2. Detection of Lymphocyte Subsets

Due to the impact of the epidemic during the early stages of this study, it should be noted that the supply of reagents was insufficient. Thus, samples from only 33 of 68 post-recovery patients were analyzed. According to the results of each measurement, CD4^+^ T cells, CD8^+^ T cells, CD19^+^ B cells, CD56^+^ NK cells and CD4^+^/ CD8^+^ T cells were divided into below normal range, within normal range and above normal range. The corresponding quantification of each lymphocyte subset, and proportions in each of the aforementioned ranges, were calculated, with the results shown in Table 1.

**TABLE 1:** Detection results of lymphocyte subsets in 2-week post-recovery patients and healthy controls (cases, %).

| Groups | Total lymphocyte |  | CD3<sup>+</sup>T cell |  | CD3<sup>+</sup>CD4<sup>+</sup> T cell |  | CD3<sup>+</sup>CD8<sup>+</sup> T cell |  | CD19<sup>+</sup> B cell |  | CD56<sup>+</sup> NK cell |  | CD4<sup>+</sup>/CD8<sup>+</sup> |  |
| --- | --- | --- | --- | --- | --- | --- | --- | --- | --- | --- | --- | --- | --- | --- |
|  | RP | HCs | RP | HCs | RP | HCs | RP | HCs | RP | HCs | RP | HCs | RP | HCs |
| Below normal values (n,%) | 22<br>(66.67) | 5<br>(17.86) | 18<br>(54.55) | 5<br>(17.86) | 18<br>(54.55) | 5<br>(17.86) | 27<br>(81.82) | 4<br>(14.29) | 23<br>(69.70) | 15<br>(53.57) | 17<br>(51.52) | 7<br>(25.00) | 0<br>(0) | 0<br>(0) |
| Within normal values (n,%) | 9<br>(27.27) | 18<br>(64.28) | 12<br>(36.36) | 19<br>(67.86) | 13<br>(39.39) | 23<br>(82.14) | 5<br>(15.15) | 17<br>(60.71) | 9<br>(27.27) | 12<br>(42.86) | 16<br>(48.48) | 17<br>(60.71) | 29<br>(87.88) | 27<br>(96.43) |
| Above normal values (n,%) | 2<br>(6.06) | 5<br>(17.86) | 3<br>(9.09) | 4<br>(14.28) | 2<br>(6.06) | 0<br>(0) | 1<br>(3.03) | 7<br>(25.00) | 1<br>(3.03) | 1<br>(3.57) | 0<br>(0) | 4<br>(14.29) | 4<br>(12.12) | 1<br>(3.57) |

RP, patients 2 weeks after recovering from COVID-19; HCs, healthy controls

Among the 33 post-recovery patients, 22 (66.67%) patients had total lymphocyte counts below the normal range, 9 (27.27%) patients were within the normal range, and 2 (6.06%) patients were above the normal range. 18 (54.55%) patients had CD3^+^ T cell counts below the normal range, 12 (36.36%) patients were within the normal range, and 3 (9.09%) patients were above the normal range. 18 (54.55%) patients had CD4^+^ T cell counts below the normal range, 13 (39.39%) patients were within the normal range, and 2 (6.06%) patients were above the normal range. 27 (81.82%) patients had CD8^+^ T cell counts below the normal range, 5 (15.15%) patients were within the normal range, and 1 (3.03%) patient was above the normal range. 23 (69.70%) patients had CD19^+^ B cell counts below the normal range, 9 (27.27%) patients were within the normal range, and 1 (3.03%) patient was above the normal range. 17 (51.52%) patients had CD56^+^ NK cell counts below the normal range, and 16 (48.48%) patients were within the normal range. 29 (87.88%) patients had CD4^+^/CD8^+^ cell ratios within the normal range, and 4 (12.12%) patients were above the range.

Among the 28 healthy controls, 5 (17.86%) patients had total lymphocyte counts below the normal range, 18 (64.28%) patients were within the normal range, and 5 (17.86%) patients were above the normal range. 5 (17.86%) patients had CD3^+^ T cell counts below the normal range, 19 (67.86%) patients were within the normal range, and 4 (14.28%) patients were above the normal range. 5 (17.86%) patients had CD4^+^ T cell counts below the normal range, and 23 (82.14%) patients were within the normal range. 4 (14.29%) patients had CD8^+^ T cell counts below the normal range, 17 (60.71%) patients were within the normal range, and 7 (25%) patients were above the normal range. 15 (53.57%) patients had CD19^+^ B cell counts below the normal range, 12 (42.86%) patients were within the normal range, and 1 (3.57%) patient was above the normal range. 7 (25%) patients had CD56^+^ NK cell counts below the normal range, 17 (60.71%) patients were within the normal range, and 4 (14.29%) patients were above the normal range. 27 (96.43%) patients had CD4^+^/CD8^+^ cell ratios within the normal range, and 1 (3.57%) patient was above normal.

### 3.3 Detection of Cytokines

Cytokines including IL-4, IL-6, IL-10, IL-17, TNF-α, and IFN-γ were divided into within the normal range and above the normal range according to various indicators. The corresponding concentration of each cytokine and proportions in each of the aforementioned ranges were calculated (Table 2).

**TABLE 2:** Detection results of cytokines in 2-week post-recovery patients and healthy controls (cases, %).

| Groups | IL-2 | | IL-4 | | IL-6 | | IL-10 | | TNF-α | | INF-γ | | IL-17 | |
| --- | --- | --- | --- | --- | --- | --- | --- | --- | --- | --- | --- | --- | --- | --- |
|  | RP | HCs | RP | HCs | RP | HCs | RP | HCs | RP | HCs | RP | HCs | RP | HCs |
| Within normal values (n,%) | 68<br>(100) | 28<br>(100) | 55<br>(80.88) | 28<br>(100) | 54<br>(79.41) | 28<br>(100) | 67<br>(98.53) | 28<br>(100) | 61<br>(89.71) | 28<br>(100) | 68<br>(100) | 28<br>(100) | 66<br>(97.06) | 28<br>(100) |
| Above normal values (n,%) | 0<br>(0) | 0<br>(0) | 13<br>(19.12) | 0<br>(0) | 14<br>(20.59) | 0<br>(0) | 1<br>(1.47) | 0<br>(0) | 7<br>(10.29) | 0<br>(0) | 0<br>(0) | 0<br>(0) | 2<br>(2.94) | 0<br>(0) |

RP, 2-week post-cured patients who have recovered from COVID-19 infection; HCs, healthy controls

Among the 68 post-recovery patients, all 68 (100%) patients had IL-2 within the normal range. 55 (80.88%) patients had IL-4 within the normal range, and 13 (19.12%) patients were above normal. 54 (79.41%) patients had IL-6 within the normal range, and 14 (20.59%) patients were above normal. 67 (98.53%) patients had IL-10 within the normal range, and 1 (1.47%) patient was above normal. 61 (89.71%) patients had TNF-α within the normal range, and 7 (10.29%) patients were above normal. All 68 (100%) patients had IFN-γ within the normal range. 66 (97.06%) patients had IL-17 within the normal range, and 2 (2.94%) patients were above normal. Among the 28 healthy controls, all subjects had IL-2, IL-4, IL-6, IL-10, TNF-α, IFN-γ and IL-17 within the normal values.

### 3.4. Comparison of lymphocyte subsets and cytokines

Interestingly, there were significant differences in the levels of total lymphocytes, CD3^+^ T cells, CD4^+^ T cells, CD8^+^ T cells, CD19^+^ B cells, CD56^+^ NK cells, IL-2, IL-4, IL-6, IL-10, TNF-α, IFN-γ and IL-17, while the CD4^+^/CD8^+^ ratio showed no significant differences between post-recovery patients and healthy controls (*P* < 0.05). These results are shown in Table 3 and Figure 1.

**TABLE 3:** Comparison of and cytokines between 2-week post-recovery patients and healthy controls.

| Groups | Lymphocyte subsets (per/μL) | | | | Groups | Cytokines(pg/mL) | | | |
| --- | --- | --- | --- | --- | --- | --- | --- | --- | --- |
|  | RP<br>(n=68) | HCS<br>(n=28) | t/Z | P |  | RP<br>(n=68) | HCS<br>(n=28) | t/Z | P |
| Total lymphocyte | 1488±718 | 2241±488 | 4.85 | <0.01 | IL-2 | 0.31(0.14,0.80) | 0.14 (0,0.14) | -4.27 | <0.01 |
| CD3<sup>+</sup>T cell | 1055±578 | 1529±389 | 3.80 | <0.01 | IL-4 | 1.27(0.70,2.56) | 0.61(0.33,0.70) | -5.09 | <0.01 |
| CD3<sup>+</sup>CD4<sup>+</sup>T cell | 545±317 | 738±161 | 3.07 | <0.01 | IL-6 | 2.98(2.11,4.47) | 1.36(0.97,1.94) | -5.23 | <0.01 |
| CD3<sup>+</sup>CD8<sup>+</sup>T cell | 314±170 | 603±212 | 5.92 | <0.01 | IL-10 | 2.01(1.52,2.47) | 1.18(0.90,1.38) | -5.35 | <0.01 |
| CD19<sup>+</sup>B cell | 129±77 | 190±74 | 3.14 | <0.01 | TNF-α | 0.92(0.67,3.00) | 0.43(0.09,0.64) | -6.25 | <0.01 |
| CD56<sup>+</sup>NK cell | 166(83,257) | 271(174,491) | -2.80 | <0.01 | INF-γ | 0.70(0.34,1.05) | 0.13(0.00,0.34) | -5.00 | <0.01 |
| CD4<sup>+</sup>/CD8<sup>+</sup> | 1.55(1.09,2.03) | 1.36±0.50 | -1.54 | 0.12 | IL-17 | 10.25±4.25 | 5.07±3.81 | -5.58 | <0.01 |

RP, 2-week post-cured patients who have recovered from COVID-19 infection; HCs, healthy controls

**FIGURE 1:**
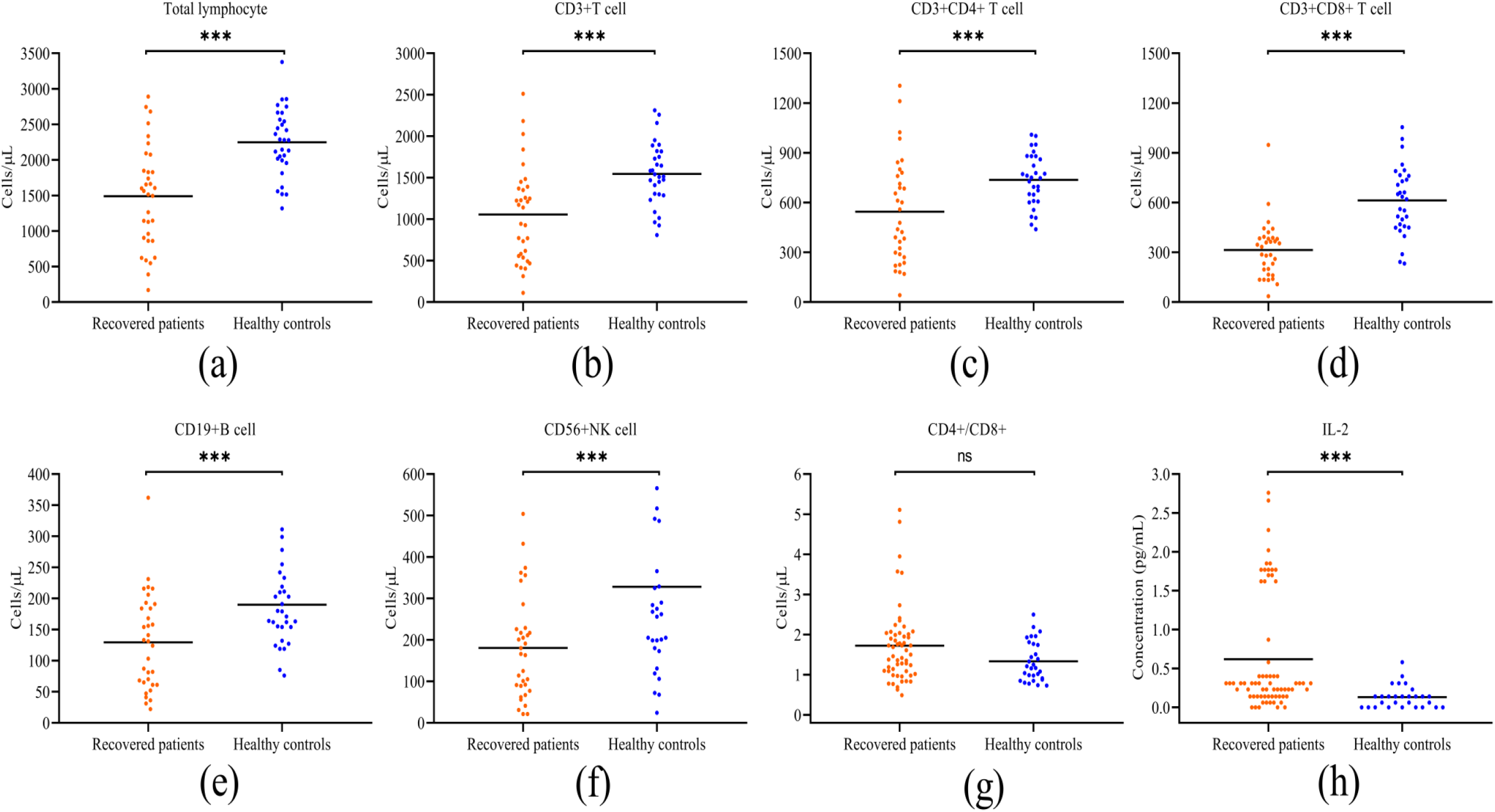
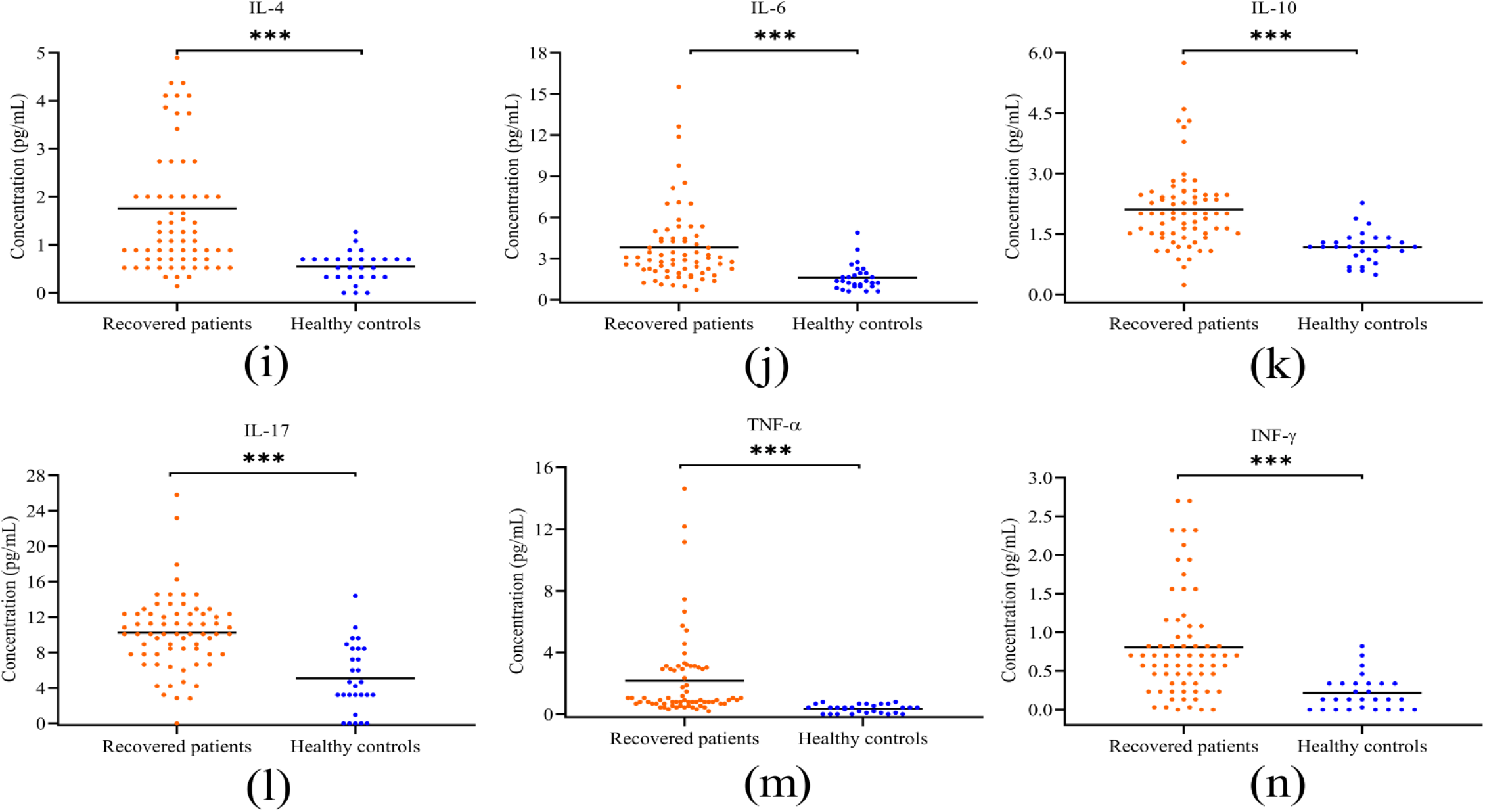
Comparison of lymphocyte subsets and cytokines between 2-week post-recovery patients and healthy controls. Individual data are presented as dots. The absolute numbers of total lymphocyte(a), CD3+ T cell (b), CD3 ^+^CD4 ^+^ T cell (c), CD3 ^+^CD8 ^+^ T cell(d), CD19 ^+^B cell(e), CD56 ^+^NK cell(f), and CD4^+^/CD8^+^(g) in the peripheral blood of recovered patients (red dot) and healthy controls (blue dot) were analyzed. The concentrations of IL-2 (h), IL-4 (i), IL-6 (j), IL-10 (k), TNF-α (l), IFN-γ(m) and IL-17(n)in the serum of recovered patients (red dot) and healthy controls (blue dot) were analyzed. ***p≤ 0.001; ns, not significant

### 3.5. Correlation between Lymphocyte Subsets and Cytokines

Considering that we observed significant differences in cytokines and lymphocyte subsets between patients who recovered from COVID-19 and healthy controls, the correlations between the cytokines and lymphocyte subsets were analyzed.

In 2-week post-recovery patients, CD3^+^ T cells were positively correlated with CD19^+^ B cells (*P* ≤ 0.001), and CD4^+^ T cells were positively correlated with CD19^+^ B cells (*P* ≤ 0.001). For cytokines, IL-2 was positively correlated with IL-4 (*P* = 0.020) and TNF-α (*P* = 0.007), IL-4 was positively correlated with IL-10 (*P* ≤ 0.001) and IL-17 (*P* = 0.032), IL-6 was positively correlated with TNF-α (*P* = 0.002), and IL-10 was positively correlated with IFN-γ (*P* = 0.001). These results are summarized in Tables 4-5, and heatmaps are shown in Figures 2-3.

**TABLE 4:** Correlation analysis between lymphocyte subsets and cytokines in 2-week post-recovery patients

| Analyses | | Cytokines(pg/mL) | | | | | | | Lymphocyte subsets (per/μL) | | | | | | |
| --- | --- | --- | --- | --- | --- | --- | --- | --- | --- | --- | --- | --- | --- | --- | --- |
| | | IL-2 | IL-4 | IL-6 | IL-10 | TNF-α | INF-γ | IL-17 | Total lymphocyte | CD3<sup>+</sup>T cell | CD3<sup>+</sup>CD4<sup>+</sup> T cell | CD3<sup>+</sup>CD8<sup>+</sup> T cell | CD19<sup>+</sup> B cell | CD56<sup>+</sup> NK cell | CD4<sup>+</sup>/CD8<sup>+</sup> |
| IL-2 | r | 1 | 0.112 | 0.056 | 0.147 | -0.155 | 0.24 | -0.095 | 0.281 | 0.299 | 0.148 | 0.38 | 0.031 | -0.069 | -0.048 |
|  | p | 0 | 0.536 | 0.757 | 0.414 | 0.39 | 0.179 | 0.6 | 0.113 | 0.092 | 0.411 | 0.029 | 0.863 | 0.703 | 0.793 |
| IL-4 | r | 0.112 | 1 | -0.254 | 0.273 | -0.074 | 0.082 | 0.628 | 0.095 | 0.164 | 0.154 | 0.175 | 0.088 | -0.109 | 0.002 |
|  | p | 0.536 | 0 | 0.154 | 0.124 | 0.684 | 0.649 | 0 | 0.599 | 0.361 | 0.393 | 0.331 | 0.627 | 0.545 | 0.991 |
| IL-6 | r | 0.056 | -0.254 | 1 | -0.081 | 0.371 | -0.119 | -0.394 | 0.13 | 0.023 | 0.123 | 0.034 | 0.219 | 0.177 | -0.001 |
|  | p | 0.757 | 0.154 | 0 | 0.655 | 0.033 | 0.509 | 0.023 | 0.47 | 0.899 | 0.496 | 0.852 | 0.221 | 0.323 | 0.994 |
| IL-10 | r | 0.147 | 0.273 | -0.081 | 1 | -0.177 | 0.04 | 0.219 | -0.012 | -0.041 | -0.168 | 0.204 | -0.088 | 0.16 | -0.242 |
|  | p | 0.414 | 0.124 | 0.655 | 0 | 0.325 | 0.827 | 0.22 | 0.949 | 0.822 | 0.349 | 0.255 | 0.627 | 0.375 | 0.175 |
| TNF-α | r | -0.155 | -0.074 | 0.371 | -0.177 | 1 | -0.322 | -0.464 | 0.106 | 0.148 | 0.112 | 0.272 | -0.149 | -0.139 | -0.104 |
|  | p | 0.39 | 0.684 | 0.033 | 0.325 | 0 | 0.067 | 0.006 | 0.558 | 0.412 | 0.534 | 0.125 | 0.409 | 0.44 | 0.563 |
| INF-γ | r | 0.24 | 0.082 | -0.119 | 0.04 | -0.322 | 1 | -0.002 | -0.231 | -0.225 | -0.191 | -0.201 | -0.194 | -0.059 | 0.09 |
|  | p | 0.179 | 0.649 | 0.509 | 0.827 | 0.067 | 0 | 0.99 | 0.197 | 0.208 | 0.287 | 0.261 | 0.279 | 0.743 | 0.617 |
| IL-17 | r | -0.095 | 0.628 | -0.394 | 0.219 | -0.464 | -0.002 | 1 | -0.088 | -0.012 | 0.088 | -0.129 | 0.04 | -0.111 | 0.124 |
|  | p | 0.6 | 0 | 0.023 | 0.22 | 0.006 | 0.99 | 0 | 0.624 | 0.949 | 0.625 | 0.474 | 0.826 | 0.539 | 0.492 |
| Total lymphocyte | r | 0.281 | 0.095 | 0.13 | -0.012 | 0.106 | -0.231 | -0.088 | 1 | 0.945 | 0.891 | 0.762 | 0.736 | 0.284 | 0.381 |
|  | p | 0.113 | 0.599 | 0.47 | 0.949 | 0.558 | 0.197 | 0.624 | 0 | 0 | 0 | 0 | 0 | 0.11 | 0.029 |
| CD3<sup>+</sup> T cell | r | 0.299 | 0.164 | 0.023 | -0.041 | 0.148 | -0.225 | -0.012 | 0.945 | 1 | 0.913 | 0.82 | 0.631 | 0.04 | 0.386 |
|  | p | 0.092 | 0.361 | 0.899 | 0.822 | 0.412 | 0.208 | 0.949 | 0 | 0 | 0 | 0 | 0 | 0.827 | 0.027 |
| CD3<sup>+</sup>CD4<sup>+</sup> T cell | r | 0.148 | 0.154 | 0.123 | -0.168 | 0.112 | -0.191 | 0.088 | 0.891 | 0.913 | 1 | 0.608 | 0.721 | 0.022 | 0.605 |
|  | p | 0.411 | 0.393 | 0.496 | 0.349 | 0.534 | 0.287 | 0.625 | 0 | 0 | 0 | 0 | 0 | 0.905 | 0 |
| CD3<sup>+</sup>CD8<sup>+</sup> T cell | r | 0.38 | 0.175 | 0.034 | 0.204 | 0.272 | -0.201 | -0.129 | 0.762 | 0.82 | 0.608 | 1 | 0.34 | 0.1 | -0.127 |
|  | p | 0.029 | 0.331 | 0.852 | 0.255 | 0.125 | 0.261 | 0.474 | 0 | 0 | 0 | 0 | 0.053 | 0.582 | 0.481 |
| CD19<sup>+</sup> B cell | r | 0.031 | 0.088 | 0.219 | -0.088 | -0.149 | -0.194 | 0.04 | 0.736 | 0.631 | 0.721 | 0.34 | 1 | 0.309 | 0.495 |
|  | p | 0.863 | 0.627 | 0.221 | 0.627 | 0.409 | 0.279 | 0.826 | 0 | 0 | 0 | 0.053 | 0 | 0.08 | 0.003 |
| CD56<sup>+</sup> NK cell | r | -0.069 | -0.109 | 0.177 | 0.16 | -0.139 | -0.059 | -0.111 | 0.284 | 0.04 | 0.022 | 0.1 | 0.309 | 1 | -0.103 |
|  | p | 0.703 | 0.545 | 0.323 | 0.375 | 0.44 | 0.743 | 0.539 | 0.11 | 0.827 | 0.905 | 0.582 | 0.08 | 0 | 0.568 |
| CD4<sup>+</sup>/CD8<sup>+</sup> | r | -0.048 | 0.002 | -0.001 | -0.242 | -0.104 | 0.09 | 0.124 | 0.381 | 0.386 | 0.605 | -0.127 | 0.495 | -0.103 | 1 |
|  | p | 0.793 | 0.991 | 0.994 | 0.175 | 0.563 | 0.617 | 0.492 | 0.029 | 0.027 | 0 | 0.481 | 0.003 | 0.568 | 0 |

**TABLE 5:**
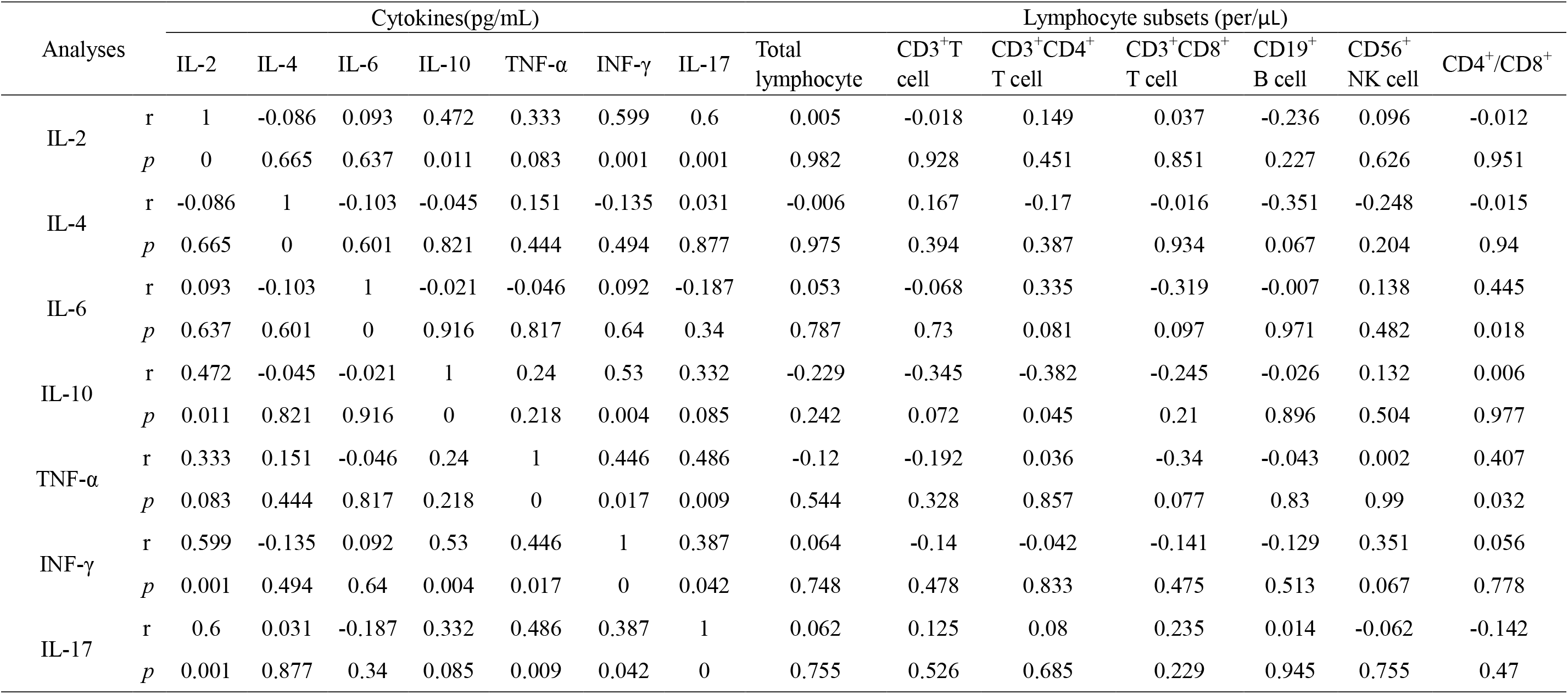
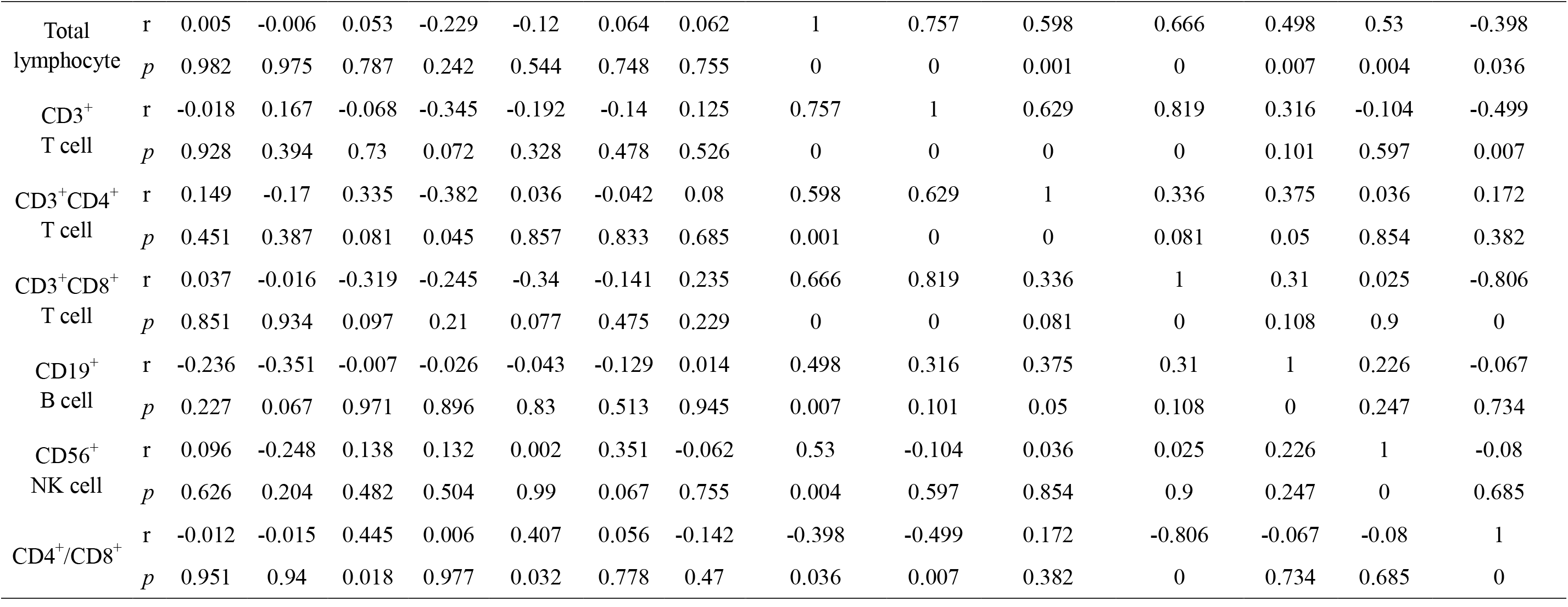
Correlation analysis between lymphocyte subsets and cytokines in healthy controls.

| Analyses | | Cytokines(pg/mL) | | | | | | | Lymphocyte subsets (per/μL) | | | | | | |
| --- | --- | --- | --- | --- | --- | --- | --- | --- | --- | --- | --- | --- | --- | --- | --- |
| | | IL-2 | IL-4 | IL-6 | IL-10 | TNF-α | INF-γ | IL-17 | Total lymphocyte | CD3<sup>+</sup>T cell | CD3<sup>+</sup>CD4<sup>+</sup> T cell | CD3<sup>+</sup>CD8<sup>+</sup> T cell | CD19<sup>+</sup> B cell | CD56<sup>+</sup> NK cell | CD4<sup>+</sup>/CD8<sup>+</sup> |
| IL-2 | r | 1 | -0.086 | 0.093 | 0.472 | 0.333 | 0.599 | 0.6 | 0.005 | -0.018 | 0.149 | 0.037 | -0.236 | 0.096 | -0.012 |
|  | p | 0 | 0.665 | 0.637 | 0.011 | 0.083 | 0.001 | 0.001 | 0.982 | 0.928 | 0.451 | 0.851 | 0.227 | 0.626 | 0.951 |
| IL-4 | r | -0.086 | 1 | -0.103 | -0.045 | 0.151 | -0.135 | 0.031 | -0.006 | 0.167 | -0.17 | -0.016 | -0.351 | -0.248 | -0.015 |
|  | p | 0.665 | 0 | 0.601 | 0.821 | 0.444 | 0.494 | 0.877 | 0.975 | 0.394 | 0.387 | 0.934 | 0.067 | 0.204 | 0.94 |
| IL-6 | r | 0.093 | -0.103 | 1 | -0.021 | -0.046 | 0.092 | -0.187 | 0.053 | -0.068 | 0.335 | -0.319 | -0.007 | 0.138 | 0.445 |
|  | p | 0.637 | 0.601 | 0 | 0.916 | 0.817 | 0.64 | 0.34 | 0.787 | 0.73 | 0.081 | 0.097 | 0.971 | 0.482 | 0.018 |
| IL-10 | r | 0.472 | -0.045 | -0.021 | 1 | 0.24 | 0.53 | 0.332 | -0.229 | -0.345 | -0.382 | -0.245 | -0.026 | 0.132 | 0.006 |
|  | p | 0.011 | 0.821 | 0.916 | 0 | 0.218 | 0.004 | 0.085 | 0.242 | 0.072 | 0.045 | 0.21 | 0.896 | 0.504 | 0.977 |
| TNF-α | r | 0.333 | 0.151 | -0.046 | 0.24 | 1 | 0.446 | 0.486 | -0.12 | -0.192 | 0.036 | -0.34 | -0.043 | 0.002 | 0.407 |
|  | p | 0.083 | 0.444 | 0.817 | 0.218 | 0 | 0.017 | 0.009 | 0.544 | 0.328 | 0.857 | 0.077 | 0.83 | 0.99 | 0.032 |
| INF-γ | r | 0.599 | -0.135 | 0.092 | 0.53 | 0.446 | 1 | 0.387 | 0.064 | -0.14 | -0.042 | -0.141 | -0.129 | 0.351 | 0.056 |
|  | p | 0.001 | 0.494 | 0.64 | 0.004 | 0.017 | 0 | 0.042 | 0.748 | 0.478 | 0.833 | 0.475 | 0.513 | 0.067 | 0.778 |
| IL-17 | r | 0.6 | 0.031 | -0.187 | 0.332 | 0.486 | 0.387 | 1 | 0.062 | 0.125 | 0.08 | 0.235 | 0.014 | -0.062 | -0.142 |
|  | p | 0.001 | 0.877 | 0.34 | 0.085 | 0.009 | 0.042 | 0 | 0.755 | 0.526 | 0.685 | 0.229 | 0.945 | 0.755 | 0.47 |
| Total lymphocyte | r | 0.005 | -0.006 | 0.053 | -0.229 | -0.12 | 0.064 | 0.062 | 1 | 0.757 | 0.598 | 0.666 | 0.498 | 0.53 | -0.398 |
|  | p | 0.982 | 0.975 | 0.787 | 0.242 | 0.544 | 0.748 | 0.755 | 0 | 0 | 0.001 | 0 | 0.007 | 0.004 | 0.036 |
| CD3<sup>+</sup> T cell | r | -0.018 | 0.167 | -0.068 | -0.345 | -0.192 | -0.14 | 0.125 | 0.757 | 1 | 0.629 | 0.819 | 0.316 | -0.104 | -0.499 |
|  | p | 0.928 | 0.394 | 0.73 | 0.072 | 0.328 | 0.478 | 0.526 | 0 | 0 | 0 | 0 | 0.101 | 0.597 | 0.007 |
| CD3<sup>+</sup>CD4<sup>+</sup> T cell | r | 0.149 | -0.17 | 0.335 | -0.382 | 0.036 | -0.042 | 0.08 | 0.598 | 0.629 | 1 | 0.336 | 0.375 | 0.036 | 0.172 |
|  | p | 0.451 | 0.387 | 0.081 | 0.045 | 0.857 | 0.833 | 0.685 | 0.001 | 0 | 0 | 0.081 | 0.05 | 0.854 | 0.382 |
| CD3<sup>+</sup>CD8<sup>+</sup> T cell | r | 0.037 | -0.016 | -0.319 | -0.245 | -0.34 | -0.141 | 0.235 | 0.666 | 0.819 | 0.336 | 1 | 0.31 | 0.025 | -0.806 |
|  | p | 0.851 | 0.934 | 0.097 | 0.21 | 0.077 | 0.475 | 0.229 | 0 | 0 | 0.081 | 0 | 0.108 | 0.9 | 0 |
| CD19<sup>+</sup> B cell | r | -0.236 | -0.351 | -0.007 | -0.026 | -0.043 | -0.129 | 0.014 | 0.498 | 0.316 | 0.375 | 0.31 | 1 | 0.226 | -0.067 |
|  | p | 0.227 | 0.067 | 0.971 | 0.896 | 0.83 | 0.513 | 0.945 | 0.007 | 0.101 | 0.05 | 0.108 | 0 | 0.247 | 0.734 |
| CD56<sup>+</sup> NK cell | r | 0.096 | -0.248 | 0.138 | 0.132 | 0.002 | 0.351 | -0.062 | 0.53 | -0.104 | 0.036 | 0.025 | 0.226 | 1 | -0.08 |
|  | p | 0.626 | 0.204 | 0.482 | 0.504 | 0.99 | 0.067 | 0.755 | 0.004 | 0.597 | 0.854 | 0.9 | 0.247 | 0 | 0.685 |
| CD4<sup>+</sup>/CD8<sup>+</sup> | r | -0.012 | -0.015 | 0.445 | 0.006 | 0.407 | 0.056 | -0.142 | -0.398 | -0.499 | 0.172 | -0.806 | -0.067 | -0.08 | 1 |
|  | p | 0.951 | 0.94 | 0.018 | 0.977 | 0.032 | 0.778 | 0.47 | 0.036 | 0.007 | 0.382 | 0 | 0.734 | 0.685 | 0 |

**FIGURE 2:**
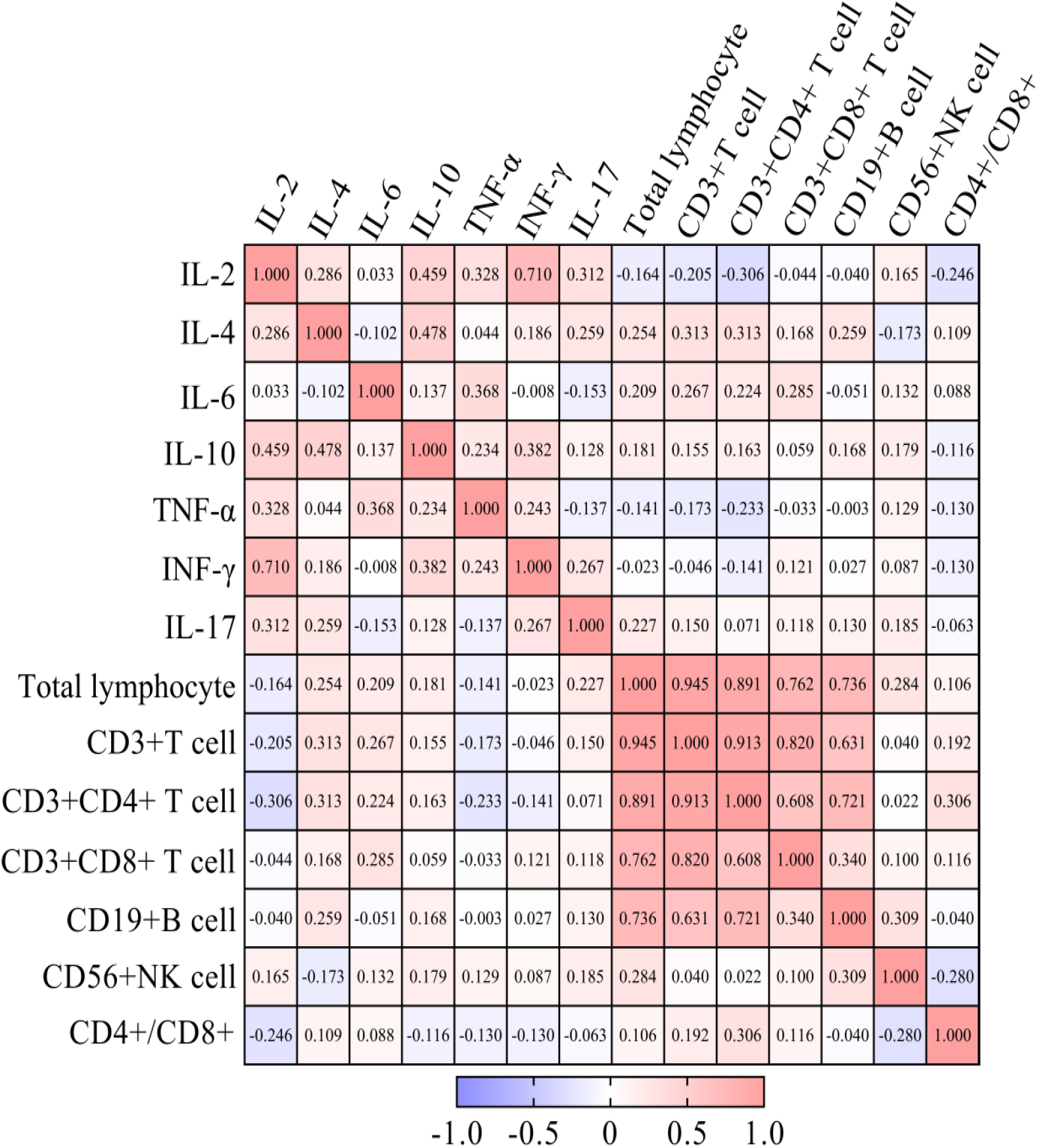
Correlation analysis was performed by R package to identify correlated variables among 2-week post-recovery patients and described by correlation heat map. The red color represents positive correlation, the blue represents negative correlation, and the number in each grid represents correlation coefficient.

**FIGURE 3:**
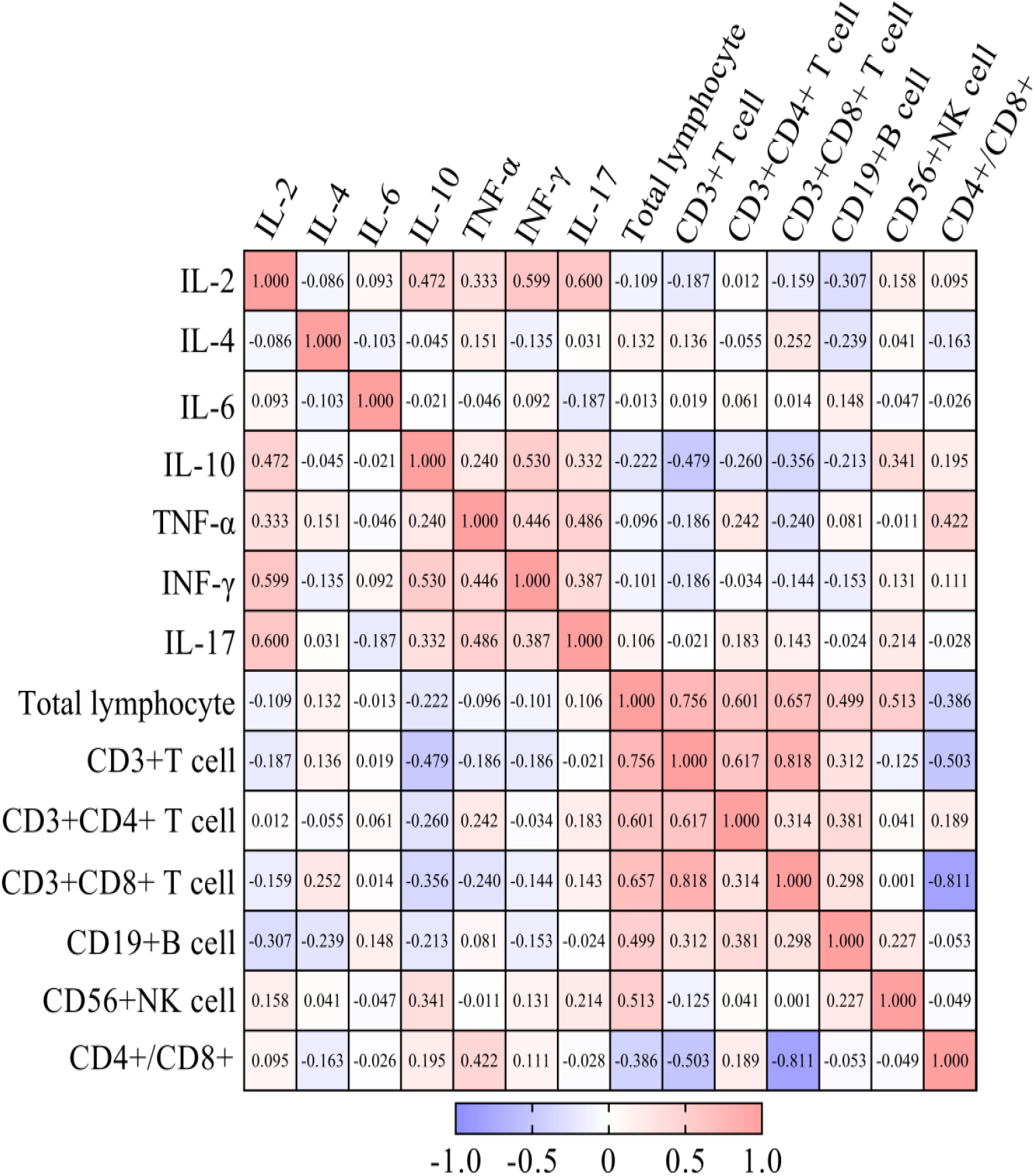
Correlation analysis was performed by R package to identify correlated variables among healthy controls and described by correlation heat map. The red color represents positive correlation, the blue represents negative correlation, and the number in each grid represents correlation coefficient.

### 3.6. ROC Curve Analysis

ROC curve analyses were conducted to evaluate the probability of changes in cytokines and peripheral lymphocyte subsets in predicting rehabilitation efficacy (Figure 4-6). The area under the ROC curve (AUC) was 0.793 (95% confidence interval (CI): 0.680-0.906) for the total lymphocyte decrease, 0.769 (95% CI: 0.649-0.890) for the CD3^+^ T cell decrease, 0.716 (95% CI: 0.585-0.848) for the CD4^+^ T cell decrease, 0.887 (95% CI: 0.798-0.976) for the CD8^+^ T cell decrease, 0.715 (95% CI: 0.588-0.843) for the CD19^+^ B cell decrease, 0.709 (95% CI: 0.579-0.840) for the CD56^+^ NK cell decrease, and 0.315 (95% CI: 0.180-0.450) for the CD4^+^/CD8^+^ ratio decrease. The area under the ROC curve (AUC) was 0.775 (95% CI: 0.667-0.874) for the IL-2 increase, 0.830 (95% CI: 0.750-0.910) for the IL-4 increase, 0.840 (95% CI: 0.754-0.927) for the IL-6 increase, 0.848 (95% CI: 0.770-0.926) for the IL-10 increase, 0.906 (95% CI: 0.849-0.963) for the TNF-α increase, 0.824 (95% CI: 0.739-0.909) for IFN-γ increase, and 0.827 (95% CI: 0.738-0.917) for the IL-17 increase. The area under the ROC curve (AUC) was 0.900 (95% CI: 0.818-0.982) for the decrease in overall peripheral lymphocyte subsets and 0.995 (95% CI: 0.985-1.000) for the overall cytokine increase. These results are shown in Figures 4-6.

**FIGURE 4:**
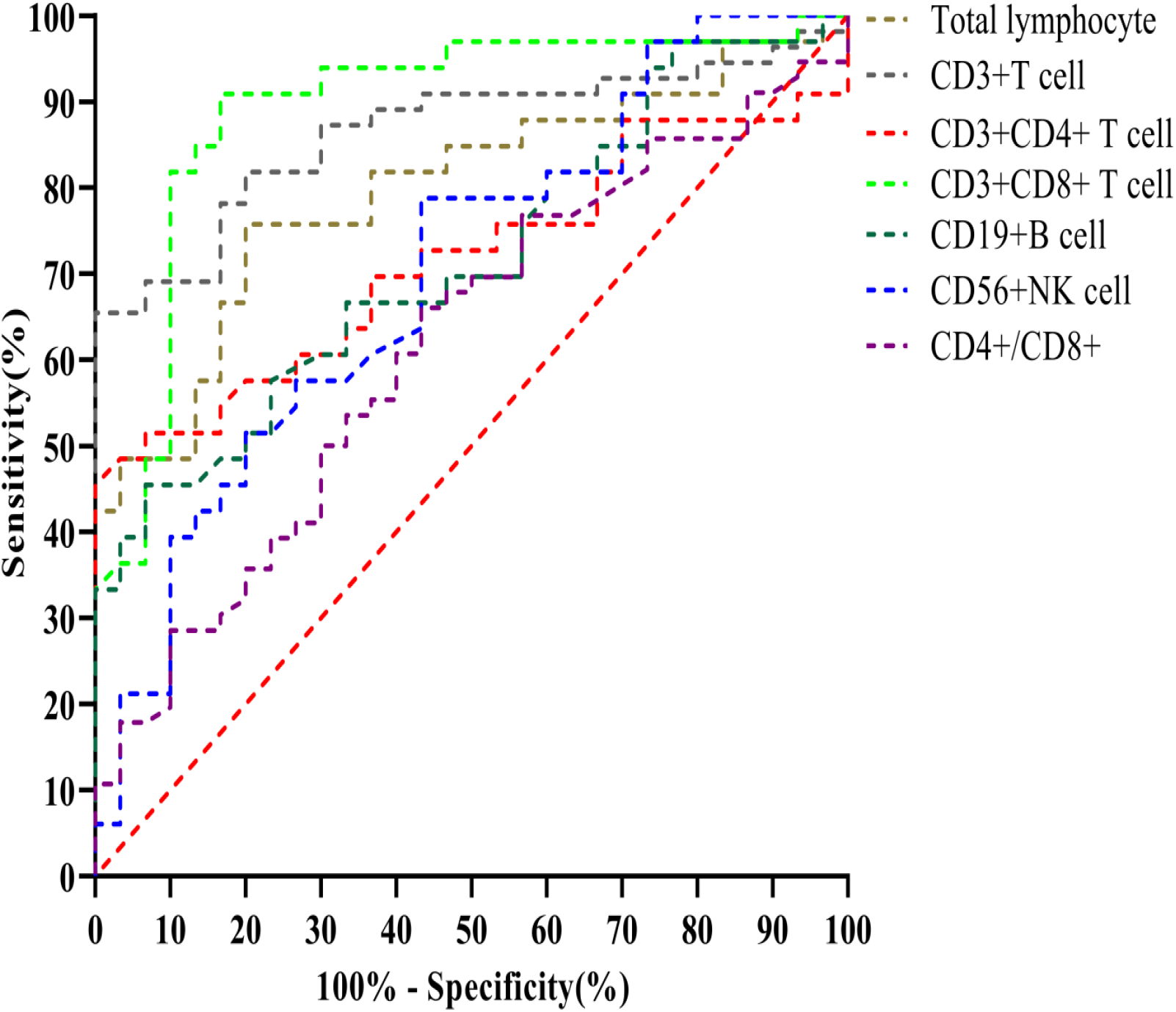
Receiver operating characteristic (ROC) curve analysis of post- recovery alteration of each lymphocyte subsets in predicting rehabilitation efficacy.

**FIGURE 5:**
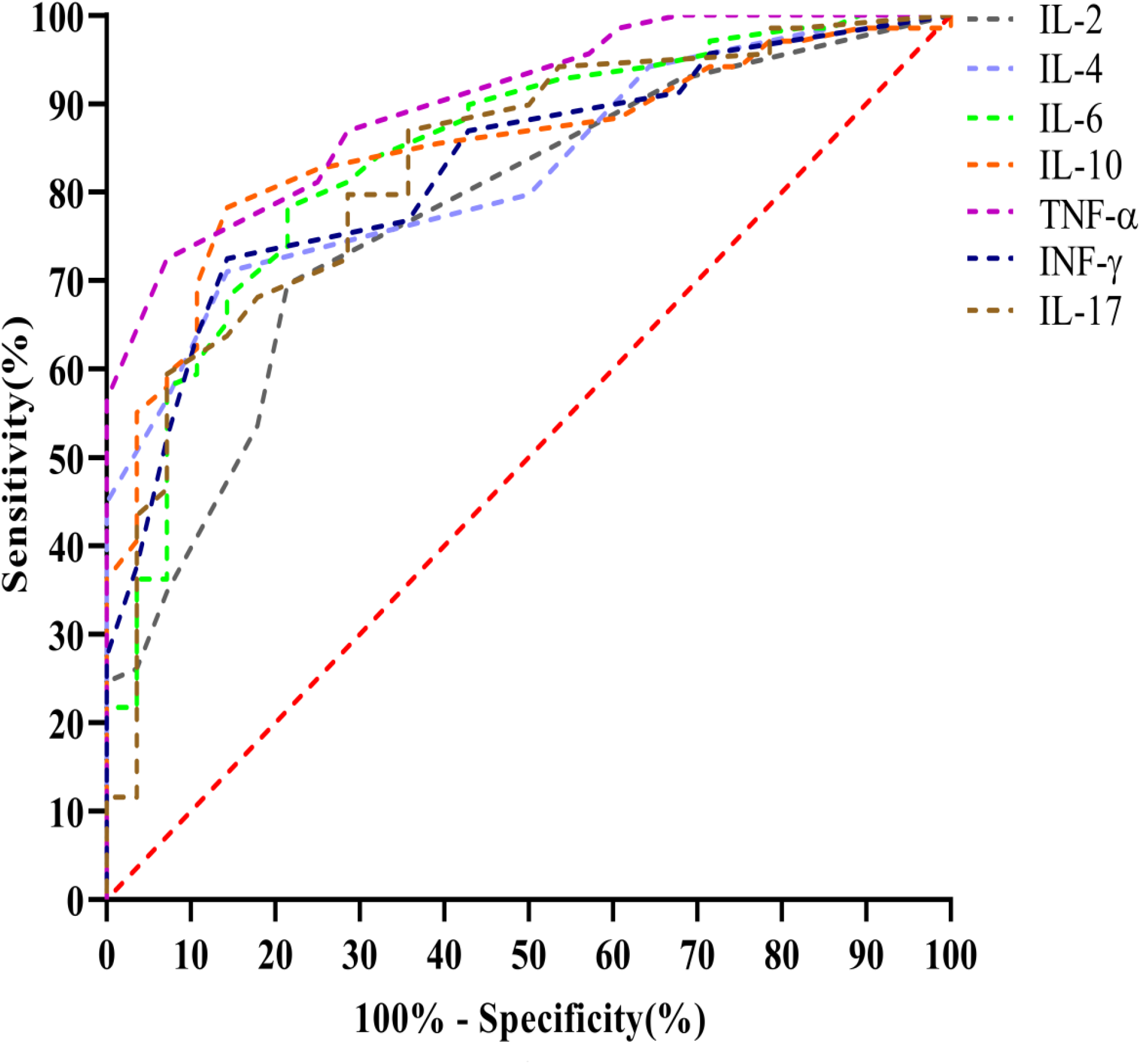
Receiver operating characteristic (ROC) curve analysis of post- recovery alteration of each cytokines in predicting rehabilitation efficacy.

**FIGURE 6:**
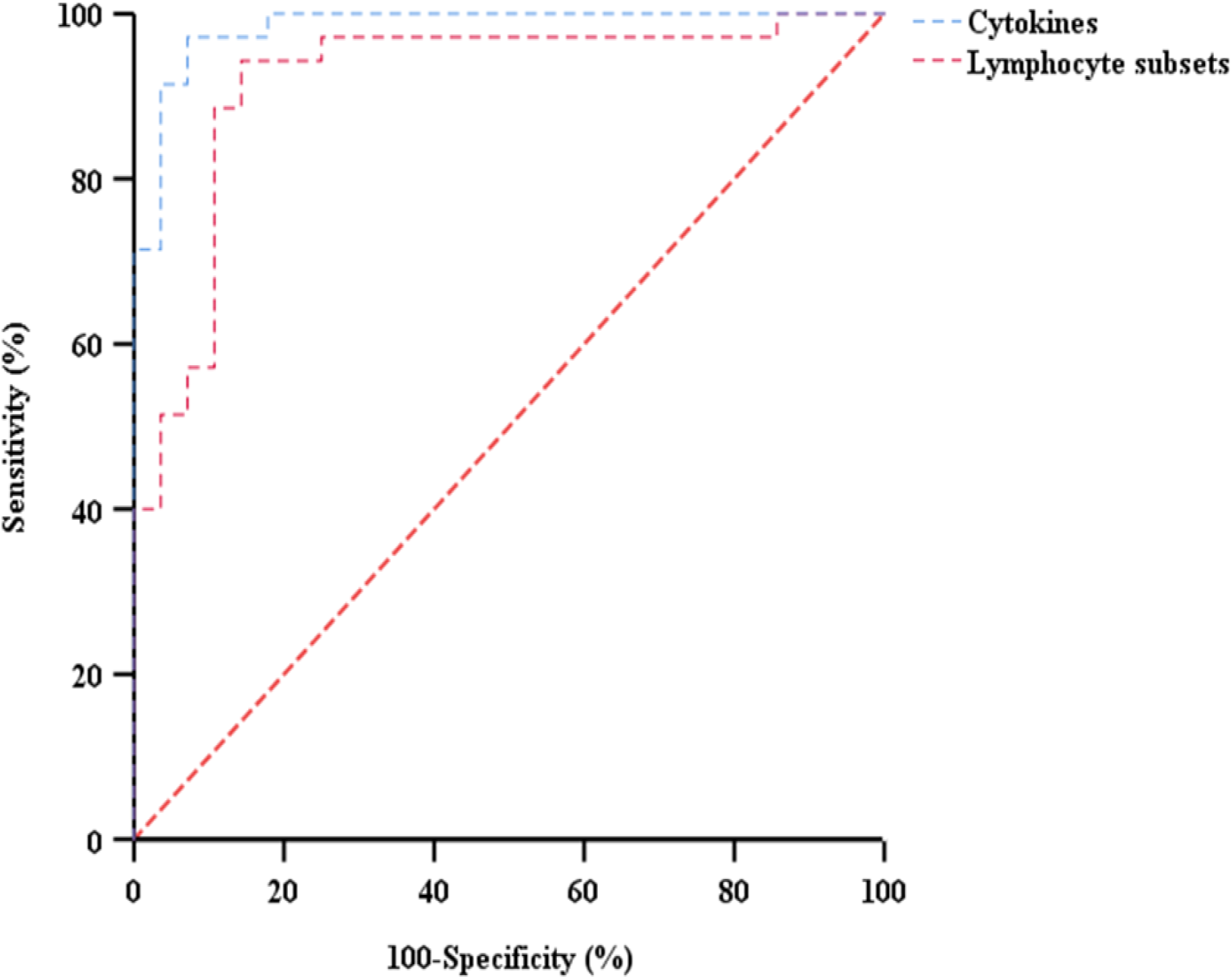
Receiver operating characteristic (ROC) curve analysis of post- recovery alteration of overall lymphocyte subsets and cytokines in predicting rehabilitation efficacy.

## 4. Discussion

The novel coronavirus is highly infectious, with the resulting disease having a poor prognosis and high mortality rate [8]. Most infected patients present with fever, fatigue, respiratory symptoms, gastrointestinal symptoms and chest imaging changes. In severe cases, the virus may lead to acute respiratory distress syndrome, septic shock, multiple organ failure, or even death [2]. It has been shown that following coronavirus infections, immune-related signaling pathways such as the toll-like

receptor and RIG-I-like receptor signaling pathways can affect the function of immune cells, such as T cells, B cells and dendritic cells, inducing the production of a large number of Th17-related cytokines, which can further lead to dysfunction of the innate and adaptive immune systems [9]. It has been also shown that targeting the microenvironment and epigenome of the infected organism to regulate relevant immune pathways are effective strategies to treat coronavirus infections [10,11,12,13]. However, the lack of knowledge regarding the impact of COVID-19 on the immune system remains a critical issue due to its rapid spread and the shortage of specific therapies [14]. In this study we characterized lymphocyte subsets and cytokines in the peripheral blood of patients 2 weeks after recovering from COVID-19 infections. Our observed results may explain why some patients fell sick after being discharged; thus, we suggest that the current criteria for hospital discharge should be re-evaluated.

Lymphocytes play critical roles in viral clearance during respiratory infections. Previous studies indicated that SARS-CoV-2 infections could lead to dysregulation of the levels of lymphocyte subsets, by showing that the absolute counts of CD3^+^ T cells, CD4^+^ T cells, CD8^+^ T cells, CD19^+^ B cells, and CD56^+^ NK cells were reduced in patients with COVID-19 [2,5]. This suggests that the coronavirus may consume many immune cells, thereby inhibiting the body’s cellular immunity. Moreover, with disease improvement, the levels of total lymphocytes, CD8^+^ T cells and B cells increased [5]. In our study, we found that in 33 post-recovery patients, most of them still exhibited lower than normal levels of total lymphocytes, total T cells, CD4^+^ T cells, CD8 ^+^ T cells, B cells, and NK cells. This is consistent with the results reported by Wen *et al*., which revealed that NK and T cells decreased in the peripheral blood of COVID-19 patients in both the early and late recovery stages [15]. Their study also demonstrated that cell-to-cell interactions may contribute to T cells and B cells, which may explain why the frequencies of peripheral blood lymphocytes gradually increased in recovering patients. In our study, compared to healthy controls, post-recovery patients displayed significantly lower absolute numbers of total lymphocytes, total T cells, CD4^+^ T cells, CD8^+^ T cells, B cells, and NK cells. This was also consistent with the above mentioned study by Wen *et al*. [15], which found that compared to healthy controls, the absolute number of CD8^+^ T cells, NK cells and CD4^+^ T cells decreased in COVID-19 patients, especially in the recovery stage; however they observed no significant changes in CD19 ^+^B cell counts, unlike in our study. Possible reasons for this discrepancy may be the direct infection of lymphocytes by SARS-CoV2, cytokine-mediated lymphocyte trafficking into infected tissues, or lymphocyte exhaustion in the peripheral blood. There may also be immune-mediated lymphocyte destruction as reported in other viral infections [15]. These potential hypotheses require further investigation. Since peripheral lymphocytes decrease following after clinical cure and discharge, it is necessary to continue to observe and follow-up with COVID-19 patients to understand the long-term effects of SARS-CoV2 on immune functions. In addition, due to the lack of clinical data in the early advanced stages of the disease, the alterations and functions of lymphocytes, particularly B cells, throughout the course of the disease need further study.

Previous studies have indicated that cytokine storms play an important role in severe COVID-19 cases [5,6]. SARS-CoV-2 binds to alveolar epithelial cells, then the virus activates the innate and adaptive immune systems, resulting in the release of a large number of cytokines. In our study, we found that in 33 post-recovery patients, most of them showed normal levels of IL-2, IL-4, IL-6, IL-10, TNF-α, IFN-γ, and IL-17. However, compared to healthy controls, patients who recovered from COVID-19 displayed significantly higher levels of IL-2, IL-4, IL-6, IL-10, TNF-α, IFN-γ and IL-17. These results suggest that SARS-CoV-2 infection, patients who recover will improve their antiviral ability within a certain period of time, then show a relatively active state of immunity. These alterations in cytokines were also found in the pneumonia caused by SARS-CoV-1. In a study by Fan *et al*., stimulation of peripheral blood mononuclear cells from patients 4 years after recovering from SARS-CoV using S peptides resulted in significantly higher levels of IFN-γ production. However, studies characterizing cytokines from patients after recovering from SARS-CoV-2 infections remain limited, and require to further investigation.

Memory T cells consist of both CD4^+^ and CD8^+^ T cells that can rapidly acquire effector functions to eliminate infected cells and secrete cytokines that inhibit replication of pathogens and regulate immune responses. After stimulation with specific antigens, memory CD4^+^ T cells differentiate into effector cells. Based on their cytokine production, CD4^+^ T cells can be classified as T-helper (Th)1 and Th2 cells. Th1 cells secrete IL-2, IFN-γ, and TNF-α, and participate in the cellular immune response. On the other hand, Th2 cells secrete IL-4, IL-6 and IL-10 and enhance humoral immune responses [16]. Among post-recovery patients, but not healthy controls, T cells, and especially CD4^+^ T cells, were positively correlated with B cell counts, and IL-2 was positively correlated with CD8^+^ T cell levels. The study by Wen *et al*. reported that in COVID-19 patients, T cell-B cell interactions induce T cells to produce IL-2, promoting the proliferation of B cells, which may explain the correlation between T cells and B cells [15]. Unlike in healthy controls that showed no correlations, among patients who recovered from COVID-19, CD4^+^ T cells were positively correlated with CD8^+^ T cell counts. Wen *et al*.’s study characterized T and NK cell responses in the blood of recovered COVID-19 patients, and showed that CD4^+^ T cells were the main participants in combating the infection, and clonally-expanded CD8^+^ T cells in the peripheral blood of help control the spread of the virus [15]. Additionally, among post-recovery patients, IL-6 was positively correlated with TNF-α and IFN-γ. This was consistent with a study by Hunter and Jones, who demonstrated that the main activators of IL-6 expression are TNF-α and IL-1β [17]. In combination with a separate study, which reported that patients with severe cases of COVID-19 showed increases in IL-6 and IFN-γ [6], we propose the hypothesis that IL-6 and IFN-γ can be independent predictors for rehabilitation efficacy. This hypothesis could provide valuable insight into the cellular immune response in patients who recovered from SARS-CoV-2, and for the design of vaccines against SARS-CoV-2.

In combination with the above findings, patients who recovered from COVID-19 infections had a concomitant significant decrease in lymphocyte subsets and increase in serum cytokines. ROC curve analyses identified the post-recovery decrease in lymphocyte subsets and the increase of cytokines as independent predictors for rehabilitation efficacy. Together, the recovery of immune function might be a reliable indicator of rehabilitation. By combining the peripheral cell counts and the secreted cytokines, we can evaluate the immune function of convalescent patients more effectively.

There are some limitations in this study. Firstly, due to the lack of clinical data during the early infection stage, continuous observational data from the same cases are absent, and potential influence by early events are not considered. Secondly, the sample size was relatively small in comparison with Wuhan where the disease originated, which may have some impact on the statistical results. In future experiments, we will conduct follow-up studies in the patients who recovered from COVID-19, and determine a quantitative basis for intervention of rehabilitation measures. This will help treat the diseases at an earlier stage by promoting medical intervention in a timely manner. Moreover, it may be beneficial to analyze if a particular population has an added immunological advantage while combating the virus.

## 5. Conclusions

In the present study, we applied flow cytometry to comprehensively characterize changes in circulating lymphocyte subsets and cytokines in patients who have recovered from COVID-19, as an assessment of their immune status following discharge. The results of both lymphocyte subsets and cytokines indicated that the immune system gradually recovers; however, the sustained hyper-inflammatory response lasting more than 14 days following discharge suggests the need for continued medical observation after patients are discharged from the hospital. Longitudinal studies of recovered patients in a larger cohort might help to better understand the consequences of the disease.

## Data Availability

The data are stored in the laboratory database.

## Data Availability

The data are stored in the laboratory database.

## Acknowledgments

This work was financially supported by Shanxi Province Social Development Program (No. 202003D31014/GZ).

## References

[1] H. Z. Lu, C. W. Stratton, Y. W. Tang, “Outbreak of pneumonia of unknown etiology in Wuhan China: the mystery and the miracle,” Journal Of Medical Virology, vol. 92, no. 4, pp. 401–402, 2020.

[2] C. L. Huang, Y. M. Wang, X. W. Li et al., “Clinical features of patients infected with 2019 novel coronavirus in Wuhan, China,” Lancet, vol. 395, no. 10223, pp. 497–506, 2020.

[3] P. F. Tang, Z. Y. Hou, X. B. Wu et al., “Expert consensus on management principles of orthopedic emergency in the epidemic of corona virus disease,” Chinese Medical Journal, vol. 133, no. 9, pp. 1096–1098, 2020.

[4] V. Balachandar, I. Mahalaxmi, M. Subramaniam et al., “Characteristics of lymphocyte subsets and cytokines in peripheral blood of 123 hospitalized,” The Science Of Total Environment, vol. 729, no. 139021, Article ID 32360909, 2020.

[5] J. L. Liu, S. M. Li, J. Liu et al., “Longitudinal characteristics of lymphocyte responses and cytokine profiles in the peripheral blood of SARS-CoV-2 infected patients,” EBioMedicine, vol. 55, no. 102763, Article ID 32361250, 2020.

[6] C. Zhang, Z. Wu, J. W. Li et al., “Cytokine release syndrome in severe COVID-19: interleukin-6 receptor antagonist tocilizumab may be the key to reduce mortality,” International Journal Of Antimicrobial Agents, vol. 55, no. 5, Article ID 32234467, 2020.

[7] J. Y. Zhao, J. Y. Yan, J. M. Qu, “Interpretation of ‘Diagnosis and Treatment Protocol for Novel Coronavirus Pneumonia (Trial Version 7)’,” Chinese Medical Journal, vol. 133, no. 11, pp. 1347–1349, 2020.

[8] N. Zhu, D. Y. Zhang, W. L. Wang et al., “A novel coronavirus from patients with pneumonia in China, 2019,” The New England Journal Of Medicine, vol. 382, no. 8, pp. 727–733, 2020.

[9] E. Kinder, V. Thiel, “To sense or not to sense viral RNA-essentials of coronavirus innate immune evasion,” Current Opinion In Microbiology, vol. 20, pp. 69-75, 2014.

[10] L. L. Dickey, C. L. Worne, J. L. Glover et al., “MicroRNA-155 enhances T cell trafficking and antiviral effector function in a model of coronavirus-induced neurologic disease,” Journal Of Neuroinflammation, vol. 13, no. 1, pp. 240, 2016.

[11] A. Schafer, R. S. Baric, “Epigenetic landscape during coronavirus infection,” Pathogens, vol. 6, no. 1, pp. 8, 2017

[12] L. Morales, J. C. Oliveros. R. Fernandez-Delgado et al., “SARS-CoV-encoded small RNAs contribute to infection-associated lung pathology,” Cell Host Microbe, vol. 21, no. 3, pp. 344–355, 2017.

[13] M. Poppe, S. Wittig. L. Jurida et al., “The NF-κB-dependent and -independent transcriptome and chromatin landscapes of human coronavirus 229E-infected cells,” Plos Pathogens, vol. 13, no. 3, e1006286, 2017.

[14] A. C. Alexandra, Y. J. Park. M. A. Alejandra Tortorici et al., “Structure, Function, and Antigenicity of the SARS-CoV-2 Spike Glycoprotein,” Cell, vol. 181, no. 2, pp. 281-292, 2020.

[15] W. Wen, W. R. Su. H. Tang et al., “Immune cell profiling of COVID-19 patients in the recovery stage by single-cell sequencing,” Cell Discovery, vol. 6, no. 31, Article ID 32377375, 2020.

[16] R. Okada, T. Kondo, F. Matsuki et al., “Phenotypic classification of human CD4+ T cell subsets and their differentiation,” International Immunology, vol. 20, no. 9, pp. 1189-1199, 2008.

[17] C. A. Hunter, S. A. Jones, “IL-6 as a keystone cytokines in health and disease,” Nature Immunology, vol. 16, no. 12, pp.448-457, 2015.

